# Virtually Delivered Psychosocial Intervention for Mothers Expecting a Baby with Congenital Heart Disease: A Proof-of-Concept Study of HEARTPrep

**DOI:** 10.64898/2026.06.03.26354861

**Authors:** Erica Sood, Kimberly S. Canter, Kamyar Arasteh, Anne E. Kazak

## Abstract

**Background:** Maternal mental health problems are common after prenatal diagnosis of congenital heart disease (CHD), with long-term implications for child and family wellbeing. HEARTPrep is a prenatal psychosocial intervention with three self-paced modules and corresponding telehealth sessions, delivered during pregnancy via mobile app to improve mental health and wellbeing for mothers expecting a baby with CHD. This proof-of-concept study evaluated the feasibility of HEARTPrep and examined maternal mental health and psychosocial functioning throughout participation.

**Methods:** Participants were mothers receiving care for a fetal CHD diagnosis within one health system. Feasibility was assessed via rates of enrollment and completion. Mothers completed 4-item PROMIS questionnaires assessing anxiety, depression, and social isolation and reported self-efficacy and hope on a weekly basis throughout HEARTPrep.

**Results:** Of 34 recruited mothers, 29 (85%) enrolled and two were subsequently not eligible (delivery prior to participation, change in fetal diagnosis), resulting in a final sample of 27 mothers. The majority (n = 22, 81%) completed all three telehealth sessions and Modules 1 (n = 22, 81%) and 2 (n = 19, 70%), with just over half (n = 14, 52%) completing Module 3 prior to delivery. Mean PROMIS depression T-scores decreased from 57.5 to 52.9, and 48% of mothers had a decrease in depression scores exceeding the meaningful change threshold (half standard deviation). The percentage of mothers reporting high self-efficacy increased from 19% to 48%.

**Conclusions:** HEARTPrep is feasible and corresponds with reduced maternal depression and increased self-efficacy, supporting proof-of-concept. A randomized controlled trial is needed to determine whether HEARTPrep improves outcomes compared to a control group.

## Introduction

Congenital heart disease (CHD) is the most common birth defect and a leading cause of infant illness and death.^1^ Infants born with complex forms of CHD require cardiac surgery and intensive care soon after birth, resulting in deviation from normative postnatal milestones and separation from parents during a critical period for attachment and development. CHD is increasingly diagnosed prenatally,^2^ facilitating specialized fetal care, informed decision making, and planning for delivery and postnatal management. Prenatal diagnosis is associated with better oxygenation, less cardiac dysfunction, and fewer neurologic sequelae.^3–5^ However, receiving a prenatal diagnosis of a life altering birth defect is often a traumatic experience for expectant parents, exerting a substantial negative impact on their mental health and wellbeing.^6–8^

Studies evaluating maternal mental health after prenatal diagnosis of CHD have revealed high rates of stress, anxiety, and depression (∼22-65%),^9,10^ approximately two to three times higher than in mothers carrying a healthy fetus.^11,12^ Maternal mental health disorders during pregnancy are known to influence in-utero brain development, postpartum mental health, maternal-infant interaction, and child developmental and behavioral trajectories.^9,13–15^ Perinatal mental health is modifiable and can be improved through psychosocial interventions.^16–20^ However, such interventions are often difficult to access and don’t target the specific stressors experienced by mothers after a prenatal diagnosis. In a prior study, 88% of mothers who received a prenatal CHD diagnosis reported that they needed, but couldn’t access, interventions targeting their unique psychosocial needs during pregnancy.^21^

HEARTPrep is a novel prenatal psychosocial intervention aimed at improving mental health and wellbeing for mothers expecting a baby with CHD.^22^ Given high rates of mental health problems and widespread unmet psychosocial needs, HEARTPrep was designed as a universal psychosocial intervention with flexible delivery of intervention components to target individual maternal/family needs, aligned with the Universal and Targeted tiers of the Pediatric Psychosocial Preventative Health Model (PPPHM).^23,24^ HEARTPrep is virtually delivered via mobile app and consists of three self-paced modules containing tools for coping and videos of parents sharing their experiences and corresponding live telehealth sessions with a psychosocial provider. HEARTPrep was initially co-designed with parents across eight health systems,^25^ with subsequent pilot testing of HEARTPrep telehealth sessions and user-centered development of HEARTPrep modules.^22,26^ These preliminary studies indicate high feasibility (90% enrollment and 70-80% completion rates) and acceptability (mean ratings of 3.5 – 3.9 on a 4-point scale). Participating mothers perceived that HEARTPrep helped them feel less distressed, less alone, more prepared, and more hopeful, with opportunities to process emotions, develop coping skills, navigate relationships, access resources, and prepare for common stressors described as among the most helpful aspects.^26^ However, maternal mental health and psychosocial functioning were not directly measured in preliminary studies of HEARTPrep.

The current study aimed to demonstrate proof-of-concept for this new model of psychosocial intervention by 1) evaluating the feasibility of HEARTPrep, including self-paced modules delivered via mobile app and live telehealth sessions, and 2) examining mental health and psychosocial functioning throughout mothers’ participation in HEARTPrep.

## Methods

### Participants

Participants were expectant mothers receiving care for a fetal diagnosis of CHD at Nemours Children’s Health. Maternal participants had the option of inviting a partner to also participate. To be eligible, expectant mothers and partners had to be 18 years of age or older and English-speaking, and the anticipated infant care plan had to include cardiac surgery in the first year of life. Mothers carrying a fetus with a genetic or congenital comorbidity were included, with the exception of those for which the anticipated life expectancy was less than one year. Mothers had to be referred for study participation by 30 weeks gestation to facilitate HEARTPrep completion prior to birth. Mothers who delivered early were included in the study until delivery. Those who no longer met inclusion criteria (e.g., change in fetal diagnosis) were removed.

### Procedures

This study was reviewed and approved by the Nemours Children’s Health Institutional Review Board (1813613) and registered at ClinicalTrials.gov (NCT05129631). The initial co-design of HEARTPrep, pilot testing of HEARTPrep telehealth sessions, and user-centered development and beta testing of HEARTPrep modules have been described previously.^22,25,26^

Recruitment spanned 20 months (beginning in July 2023), with a target sample size of 20 or more participating mothers, typically sufficient to demonstrate proof-of-concept.^27^ Perinatal nurse coordinators provided eligible expectant mothers with information about the study during fetal cardiology appointments or by phone and directly referred mothers to the research team.

Expectant mothers were contacted by a research team member to conduct the informed consent process and offer the option of partner participation. Informed consent for both mothers and partners was documented electronically via DocuSign.

After consenting, expectant mothers were provided instructions on downloading the HEARTPrep mobile app and scheduled for their first telehealth session. Telehealth sessions (∼45–60 minutes) were conducted individually with each family (expectant mother or mother/partner dyad) by a licensed clinical psychologist embedded within the Nemours Cardiac Center. Expectant mothers and partners (when applicable) could participate from anywhere, with dyads together in the same room or in separate locations. After each session, participants were provided access to sequential HEARTPrep modules (three total) within the mobile app containing recorded videos of other parents sharing experiences and articles to normalize experiences and promote adaptive coping (Supplementary Table 1). Three telehealth sessions corresponding with the three modules were required for intervention completion, with an anticipated duration of approximately six weeks from HEARTPrep initiation to completion. However, flexibility was prioritized when determining the number (between 3 and 6), spacing (e.g., weekly, every other week, monthly), and timing (e.g., Monday through Friday during business hours or evenings) of telehealth sessions to meet the individual needs and preferences of families.

Maternal participants were asked to complete a 20-item questionnaire within the mobile app to assess their mental health and psychosocial functioning on a weekly basis throughout participation in HEARTPrep, starting on the day of the first telehealth session and first module assignment. Push notification reminders were sent from the app 7 days after the date of the last questionnaire completion. Assessment results were discussed during telehealth sessions to inform session content (e.g., prioritization of intervention components). Although partners could participate in HEARTPrep with the mother, they were not asked to complete questionnaires for this proof-of-concept study, as the primary goal of HEARTPrep is to improve the mental health and wellbeing of mothers expecting a baby with CHD. Both mothers and partners were paid $10 per completed telehealth session for their time and mothers were paid $10 per completed questionnaire.

### Measures

#### Clinical and Demographic Characteristics

Maternal and fetal characteristics (e.g., age, race, ethnicity, timing and type of fetal cardiac diagnosis, and comorbidities), were extracted from the electronic medical record. Area Deprivation Index (ADI; national percentile rankings from 1 [lowest level of disadvantage] to 100 [highest level of disadvantage]) was determined based on street address.^28^

#### Intervention Uptake and Completion

Rates of enrollment and completion (3 telehealth sessions, 3 modules) were tracked as measures of feasibility. Reasons for non-enrollment in the study and non-completion of telehealth sessions were also tracked.

#### Anxiety, Depression and Social Isolation

Short questionnaires (4 items each) were created from the validated PROMIS item banks for anxiety, depression, and social isolation^29,30^ to reflect symptoms and experiences thought to be most relevant for expectant mothers after a prenatal diagnosis of CHD (Supplemental File 1). Items were rated on a 5-point scale ranging from “Never” (1) to “Always” (5) and referenced the “past 7 days.” The process of identifying these outcomes of interest and creating the custom short forms in partnership with parents who had received a prenatal CHD diagnosis has been previously described.^25^ PROMIS item banks are created in accordance with rigorous scientific standards,^31^ and any subset of items from a single PROMIS item bank can be administered as a short form and converted to T-scores (mean of 50, standard deviation [SD] of 10) via the HealthMeasures Scoring Service.^32^ T-scores between 60-69 are moderately elevated and T-scores 70 and above are severely elevated.^33^ Estimates of meaningful change thresholds for adult PROMIS measures of emotional wellbeing differ across studies and populations, generally ranging from 2.3-4.5 points on a T-score metric.^34,35^ In accordance with HealthMeasures guidance for estimating meaningful change thresholds for populations in which there is no empirical literature, we used a conservative threshold of a half standard deviation (5 points on a T-score metric).^36^

#### Self-Efficacy and Hope

Brief 4-item questionnaires of self-efficacy and hope were created by the study team, in partnership with parents who had received a prenatal CHD diagnosis. These measures were informed by the PROMIS item banks for General Self-Efficacy and Meaning and Purpose,^37,38^ but items were modified to reflect the experiences relevant for expectant parents after a prenatal diagnosis of CHD (Supplemental Table 1). The decision to create new measures of self-efficacy and hope was based on 1) our prior research with parents who had received a prenatal CHD diagnosis indicating that these outcomes are of great importance, 2) our goal of identifying increased positive outcomes in addition to decreased negative outcomes, and 3) the lack of existing measures or validated item banks applicable to the experiences of self-efficacy and hope in pregnant women expecting a baby with a life-altering birth defect. Items were rated on a 5-point scale ranging from “Not At All” (1) to “Very Much” (5) and were summed to create a total raw score ranging from 4 to 20. Internal consistency (Cronbach’s α) was excellent (Self-efficacy α = 0.92, Hope α = 0.92). For the purposes of this study, high levels of self-efficacy and hope were defined as scores ≥ 16, reflecting ratings of “Quite a Bit” and “Very Much.”

### Analysis

Feasibility was evaluated using descriptive statistics (e.g., percent enrolled/completed). Potential differences in demographic and clinical characteristics between completers and non-completers were explored using chi-square tests. Maternal mental health and psychosocial functioning throughout participation in HEARTPrep was descriptively examined in four ways: 1) weekly mean scores over six weeks of HEARTPrep participation, 2) mean scores at the first and last assessments, 3) percentages of mothers scoring in the moderate and severe ranges for anxiety, depression, and social isolation at the first and last assessments, and 4) percentages of mothers whose anxiety, depression, and social isolation scores from their first to last assessment exceeded the meaningful change threshold (half standard deviation). Analyses included mothers who participated in HEARTPrep, even if they did not complete all telehealth sessions or modules.

A six-week period was selected for the descriptive analysis of weekly mean scores based on the median time from first to last HEARTPrep telehealth session in this sample (41.5 days). Due to variability in the exact timing of assessment completion, weeks were defined as 0 (assessments completed 0-7 days from HEARTPrep initiation), 1 (8-14 days), 2 (15-21 days), 3 (22-28 days), 4 (29-35 days), 5 (36-42 days), and 6 (43-49 days). The first and last assessments were defined as the assessment completed on the day of or closest to HEARTPrep initiation (i.e., the date of the first HEARTPrep telehealth session and first Module assignment) and the assessment completed on the day of or closest to HEARTPrep completion (or last telehealth session for non-completers), respectively. Descriptive analyses involving the first and last assessments excluded participants who completed only one single assessment. Inferential statistics were not used to evaluate changes in maternal mental health and psychosocial functioning, in line with standards for small pilot studies^39^ and with the study aim to demonstrate proof-of-concept for this new model of psychosocial intervention.

## Results

### Intervention Uptake

Thirty-four expectant mothers were referred for study participation, and 29 (85%) enrolled. Of the five who didn’t enroll, three were unable to be reached, one expressed that she had too many competing demands, and one indicated already feeling prepared due to a family history of CHD. One enrolled mother was later removed from the study due to change in fetal diagnosis no longer meeting eligibility criteria and another delivered early prior to starting HEARTPrep, resulting in 27 eligible enrolled mothers (Figure 1).

**Figure 1.**
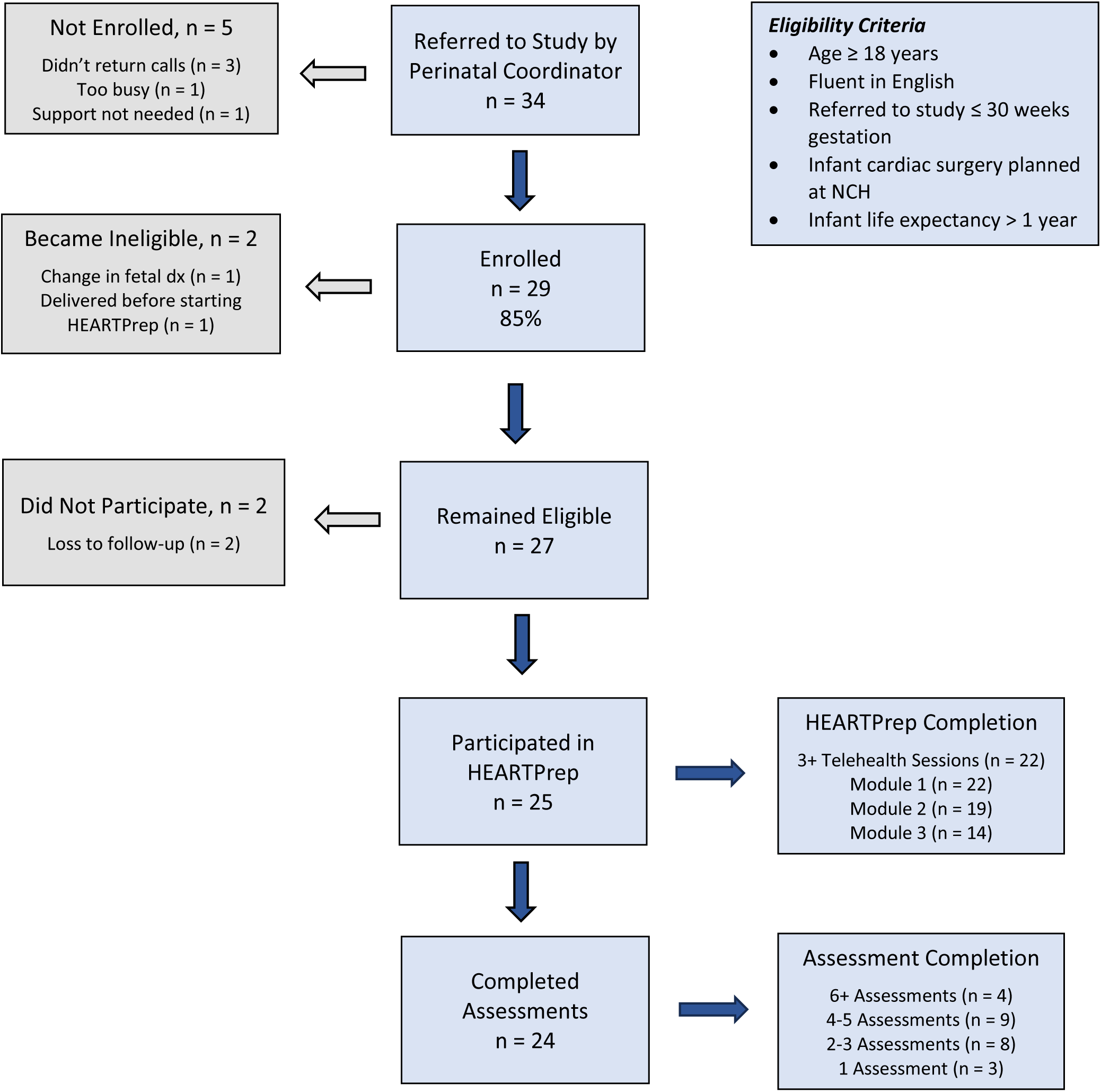
Rates of Enrollment and Completion

Maternal participants were diverse with regard to age (18 – 43 years), neighborhood socioeconomic disadvantage (ADI 16 to 97), insurance type (44% Medicaid), gestational age at first fetal cardiology consultation (15 – 28 weeks), and fetal cardiac diagnosis (Table 1). At enrollment, mean gestational age was 28 ± 3 weeks (range 22 – 34), corresponding with a mean of 48 ± 25 days (range 11 - 114) from first fetal cardiology consultation.

**Table 1.** Maternal and Fetal Characteristics (N = 27)

| **Maternal/Fetal Characteristic** | **N (%) / Mean ± SD** |
| --- | --- |
| Maternal Age (Years) <sup>a</sup> |  |
| 18-24 | 3 (11%) |
| 25-29 | 10 (37%) |
| 30-34 | 11 (41%) |
| 35-43 | 3 (11%) |
| Maternal Race/Ethnicity |  |
| Non-Hispanic White | 18 (67%) |
| Non-Hispanic Black | 9 (33%) |
| Maternal Insurance Type |  |
| Private | 15 (56%) |
| Medicaid | 12 (44%) |
| Area Deprivation Index <sup>b</sup> | 55.3 ± 21.8 |
| Gestational Age at First Fetal Cardiology Consultation |  |
| 15-19 weeks | 8 (30%) |
| 20-24 weeks | 15 (56%) |
| 25-28 weeks | 4 (15%) |
| Fetal Cardiac Diagnosis |  |
| Single-Ventricle Physiology | 9 (33%) |
| Biventricular Physiology | 18 (67%) |
| Comorbid Fetal Diagnosis |  |
| Down Syndrome | 3 (11%) |
| Comorbid Congenital Defect <sup>c</sup> | 1 (4%) |

<sup>a</sup> At time of enrollment
<sup>b</sup> Area Deprivation Index reported as mean percentile ± standard deviation. Higher percentiles reflect a higher level of disadvantage.
<sup>c</sup> Comorbid congenital defect requiring surgery during infancy.

Sixteen partners enrolled, corresponding with 59% of eligible enrolled mothers. In cases where the expectant mother had a partner who did not participate, the most common reasons provided were work schedules or competing demands.

### Intervention Completion

Twenty-five of the 27 eligible enrolled mothers (93%) participated in HEARTPrep (two were lost to follow-up and didn’t complete any telehealth sessions or modules). Completion of HEARTPrep telehealth sessions and modules are reported separately below.

#### HEARTPrep Telehealth Sessions

Twenty-two mothers (81% of the 27 eligible) completed the three sessions required for intervention completion (Figure 1). Reasons for non-completion were complications resulting in maternal hospitalization (N = 1), early delivery (N = 1), and perceived lack of need for additional psychosocial intervention (N = 1). All three mothers who participated but didn’t complete were carrying a fetus with Down Syndrome or a comorbid congenital defect. Among completers, time from the first to last session ranged from 16 to 93 days (median = 41.5), with 10 mothers choosing to attend more than the minimum of three sessions required for intervention completion (9 attended 4 sessions, 1 attended 6 sessions).

#### HEARTPrep Modules

Twenty-two mothers (81% of the 27 eligible) completed Module 1, 19 (70%) completed Module 2, and 14 (52%) completed Module 3. Demographic characteristics did not significantly differ between those who completed all three modules versus those who did not (43% vs. 46% Medicaid, 43% vs. 23% Black, mean age 30.6 vs. 28.6, mean ADI 56.4 vs. 54.2). However, mothers who didn’t complete all three modules were significantly more likely to be carrying a fetus with Down Syndrome or a comorbid congenital defect (31% vs. 0%, p = 0.04). While reasons for non-completion of HEARTPrep modules were not systematically collected, some mothers described competing demands interfering with completion of self-paced modules, particularly as they approached the delivery date, and a couple mothers noted that they didn’t feel emotionally ready or that it would be beneficial to watch videos of other parents sharing their experiences.

### Anxiety, Depression, and Social Isolation

Of the 25 mothers who participated in HEARTPrep, 24 completed one or more assessments (Figure 1; median of 4 assessments per participant), averaging 1 for approximately every 10 days of HEARTPrep. Figure 2 displays weekly mean scores over six weeks of HEARTPrep participation. Given that some participants completed HEARTPrep prior to six weeks and no participants completed assessments at both 5 and 6 weeks, assessments from weeks 5 and 6 were combined. Mean scores fluctuated from week to week with variation in the number of participants at each weekly time point. Nevertheless, depression showed a downward trend, with mean scores at 5-6 weeks more than half a standard deviation lower than week 0. Anxiety and social isolation also evidenced slight downward trends, approaching the threshold of half a standard deviation difference from week 0 to weeks 5-6. Of note, two participants who continued to engage in HEARTPrep well past the median number of days (65 and 93 days) continued to report elevated levels of anxiety and depression throughout and beyond the six-week period. When these two participants were removed, mean scores were lower but the general trajectories were unchanged (Depression: 56.8 at week 0, 49.7 at weeks 5-6; Anxiety: 58.1 at week 0, 53.8 at weeks 5-6).

**Figure 2.**
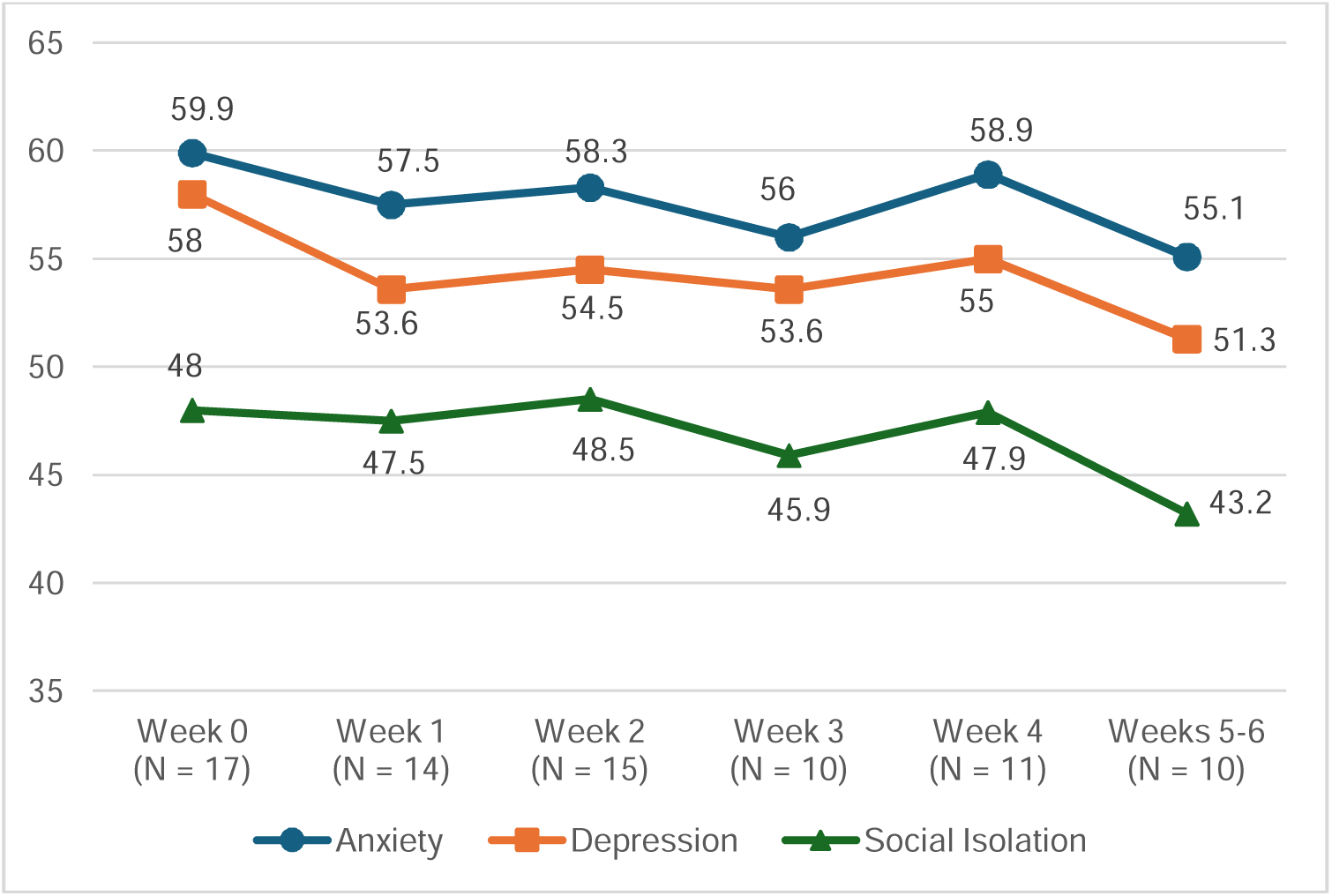
Mean Scores Over Six Weeks of HEARTPrep Participation: Anxiety, Depression and Social Isolation

At the first assessment, 12 participants (57%) were elevated on anxiety (9 moderate, 3 severe), 7 (33%) on depression (5 moderate, 2 severe), and 2 (10%) on social isolation (2 moderate) (excluding three participants who completed only one single assessment; N= 21). Mean depression scores decreased from 57.5 at the first assessment to 52.9 at the last assessment, whereas mean anxiety and social isolation scores were relatively unchanged from 60.5 to 59.3 and 47.0 to 46.2, respectively. Ten (48%) participants had a decrease in depression scores that exceeded the meaningful change threshold (half standard deviation) and 1 (5%) had an increase. Six (29%) participants had a decrease in anxiety scores that exceeded the meaningful change threshold and 4 (19%) had an increase (half of these were from the lowest possible score at baseline to a higher score within the normal range). For social isolation, 5 (24%) had a decrease in scores that exceeded the meaningful change threshold and 2 (10%) had an increase. At the last assessment, 9 participants (43%) were elevated on anxiety (8 moderate, 1 severe), 4 (19%) on depression (3 moderate, 1 severe), and 3 (14%) on social isolation (3 moderate).

### Self-efficacy and Hope

Both self-efficacy and hope demonstrated upward trajectories over 6 weeks of HEARTPrep participation (Figure 3). While many participants (12; 57%) reported high levels of hope at the time of their first assessment (scores ≥16), levels of self-efficacy were lower, with only 4 (19%) participants reporting high levels. Mean self-efficacy scores increased from 13.1 at the first assessment to 14.7 at the last assessment and mean hope scores increased from 15.5 to 16.9. At the last assessment, 14 participants (67%) reported high levels of hope and 10 (48%) reported high levels of self-efficacy.

**Figure 3.**
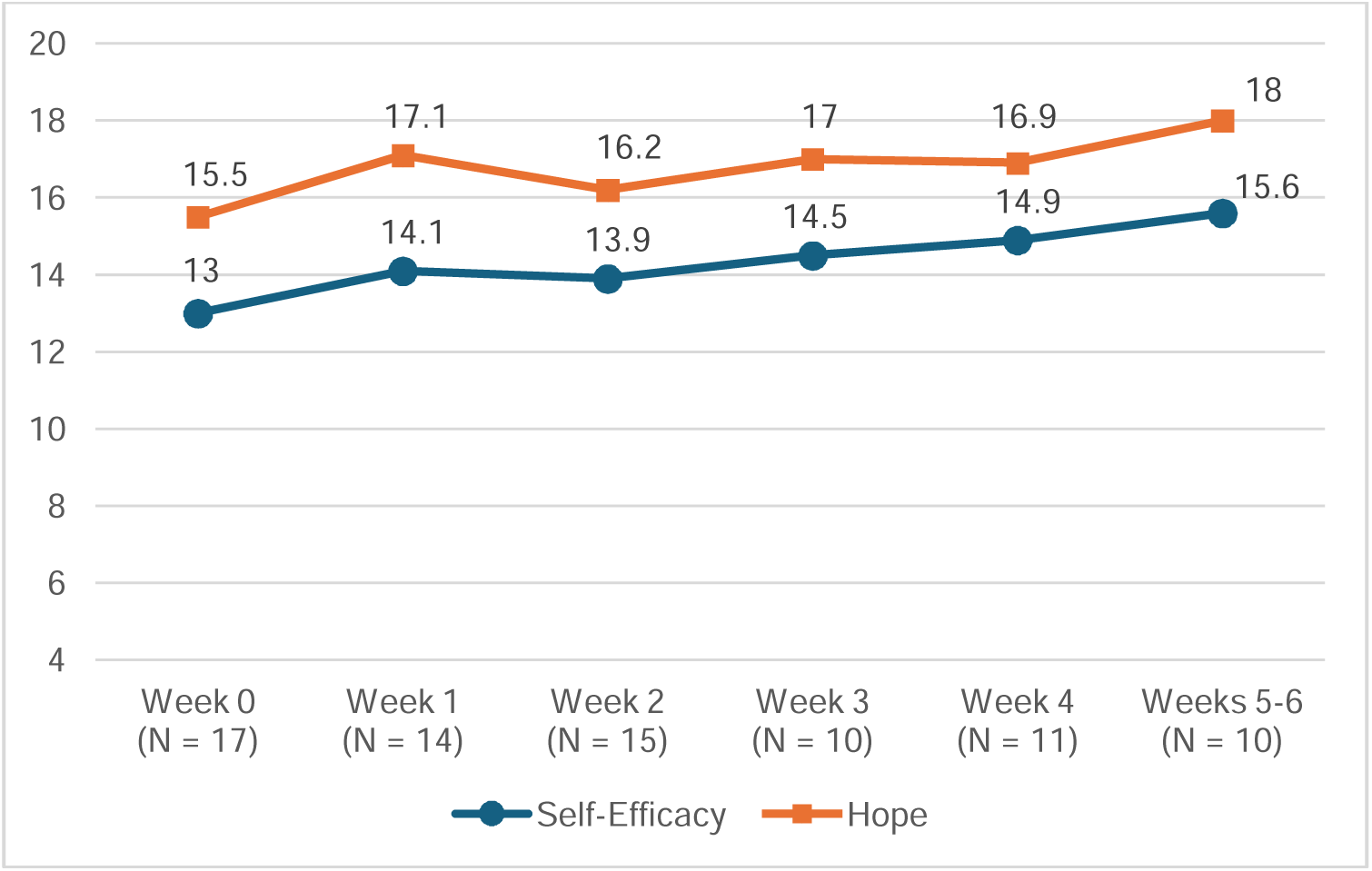
Mean Scores Over Six Weeks of HEARTPrep Participation: Self-Efficacy and Hope

## Discussion

This study builds on prior evidence for the feasibility of psychosocial intervention during pregnancy after prenatal diagnosis of CHD. High rates of HEARTPrep uptake are consistent with existing evidence that expectant parents want psychosocial intervention after prenatal diagnosis and will choose to participate when the introduction is introduced by their fetal care team and delivered in an accessible manner.^21,25,40^ Results also underscore the importance of flexibility in psychosocial intervention delivery based on individual needs/preferences and other factors including gestational age at diagnosis. Even with flexibility, competing demands and early delivery negatively impacted completion of HEARTPrep modules and telehealth sessions, highlighting the unique challenges of structured psychosocial intervention during an intense and time-limited period.

High rates of clinically elevated maternal anxiety (57%) and depression (33%) at the first assessment support the need for targeted psychosocial intervention during pregnancy. With HEARTPrep participation, half of participants experienced an improvement in depression scores considered to be clinically meaningful, and the percentage of participants who scored in the clinically elevated range on depression decreased to a rate similar to that of mothers carrying a healthy fetus.^41^ These results provide proof-of-concept for this new model of prenatal psychosocial intervention. Prenatal depression is the strongest predictor of postpartum depression,^42^ and both are established risk factors for impaired maternal-infant interaction (e.g., lower maternal sensitivity/responsiveness), maternal and child health problems, and child developmental disabilities.^14,15,43,44^ There were also promising trends in self-efficacy with HEARTPrep participation. Despite its inclusion in conceptual models focused on parental mental health,^45^ few empirical studies in the CHD literature have included direct measurement of parental self-efficacy. However, research in other populations indicates that parental self-efficacy can influence parent-child relationships, parental mental health outcomes, and child development, making it an important area for future research.^46–48^

It is notable that 43% of mothers continued to report elevated anxiety with HEARTPrep participation. The weeks leading up the birth of a baby with a life-threatening condition are by nature an anxiety-provoking time, and anxiety could be more challenging to ameliorate during this time. As noted previously, two participants who remained engaged in HEARTPrep well past the median number of days continued to report elevated levels of anxiety and depression. More time or sessions in HEARTPrep beyond the three sessions required for intervention completion did not appear to mitigate these symptoms. HEARTPrep was designed as a brief, feasible, and accessible intervention to promote the mental health and wellbeing of all mothers expecting a baby with CHD, with flexibility in the number, timing, and content of telehealth sessions to target individual maternal/family needs. The PPPHM is a three-tier model of family psychosocial risk used to guide intervention approaches in pediatric healthcare settings.^23^ Based on a recent scoping review of the pediatric healthcare literature,^24^ the majority of families fall within the Universal (∼55%) and Targeted (∼34%) ranges, requiring universal family-centered preparation and support and targeted interventions specific to family needs, while a minority of families (∼11%) fall within the Clinical Range, requiring more intensive mental health and psychosocial services. Mothers with persistent clinically elevated anxiety and depression likely require established evidence-based treatments for mental health disorders.

Surprisingly, rates of clinically elevated social isolation were quite low in this sample, in contrast with numerous qualitative studies suggesting that expectant mothers often feel lonely and isolated after prenatal diagnosis.^25,49^ Examinations of raw to T-score conversions for PROMIS Social Isolation short forms indicate that higher raw scores are typically required to reach the thresholds for moderate and severe clinical elevations when compared to the short forms for Anxiety and Depression,^50^ suggesting that some degree of social isolation may be considered normative. In the case of prenatal diagnosis, feelings of loneliness and social isolation are common reactions to a stressful and isolating experience among individuals who may otherwise feel socially connected, differing substantially from long-standing, pervasive social isolation. These results suggest that a different measure or set of norms may be needed to identify feelings of loneliness and social isolation specific to the experience of prenatal diagnosis.

As this was a proof-of-concept study with no control group, it is not known how scores would have changed without participation in HEARTPrep. Most published studies of maternal mental health after prenatal diagnosis of CHD are cross-sectional, precluding conclusions regarding the natural trajectories of these symptoms over time. Upon starting HEARTPrep, participants were at varying points in their pregnancy (range 22-34 weeks) with varying time since the prenatal diagnosis (range 11-114 days), so changes during HEARTPrep participation are not likely to simply reflect initial adjustment to the prenatal diagnosis or experiences tied to a specific point in pregnancy. Studies in the general population indicate that mental health disorders typically remain the same or worsen over the course of pregnancy and don’t usually improve on their own without intervention.^51–55^ However, mothers carrying a fetus with CHD are provided with ongoing preparation and support from a multidisciplinary healthcare team and it is possible that patterns observed in the general population don’t apply to this group of mothers.

Longitudinal research on the trajectories of mental health symptoms following prenatal diagnosis is needed. Additionally, randomized clinical trials of prenatal psychosocial interventions, including HEARTPrep, will determine whether psychosocial intervention can significantly and meaningfully improve outcomes when compared to a control group.

While not a specific aim of this study, results provide evidence for the feasibility of app-based mental health and psychosocial screening throughout the pregnancy after prenatal diagnosis. Anecdotally, many mothers commented on the ease of completing PROMIS short forms through the mobile app and some noted that they appreciated the opportunity to check in on their own wellbeing on a consistent basis. Of note, these assessments were conducted within the context of a structured psychosocial support program through which mothers knew they would have the opportunity to discuss their mental health and wellbeing. Future work could examine the feasibility and utility of weekly mental health and psychosocial screening via mobile app for mothers expecting a baby with CHD outside of a structured psychosocial intervention.

Several limitations should be considered. There was variability in the timing of questionnaire completion, resulting in missing data for the weekly analysis and a range in timing of the first and last assessments. Additionally, questionnaires started at the time of HEARTPrep initiation and therefore mental health trajectories prior to this point are not known. There were likely multiple factors (e.g., specific stressors and supports) that contributed to fluctuations in symptom trajectories over time. As previously noted, without a control group, it is not possible to determine whether improvements are attributable to HEARTPrep versus these other factors.

Participants were English speaking, with a notable lack of representation from Hispanic/Latina mothers. Inclusion criteria included English language and referral for study participation by 30 weeks gestation, limiting sample diversity, particularly when considering the known disparities in the presence and timing of prenatal diagnosis affecting Hispanic families.^56,57^ A Spanish-language version of HEARTPrep has since been developed and will be tested in future studies. Additionally, research should examine models of brief psychosocial intervention for mothers receiving a prenatal diagnosis later in the third trimester.

In conclusion, this study provides proof-of-concept for HEARTPrep, a virtually-delivered prenatal psychosocial intervention for mothers expecting a baby with CHD and their partners. HEARTPrep demonstrated feasibility and corresponded with reduced maternal depression and increased maternal self-efficacy. A future randomized controlled trial with standardized assessment timepoints, while also balancing the flexibility necessary for psychosocial intervention during pregnancy after prenatal diagnosis, is needed to determine whether HEARTPrep significantly and meaningfully improves outcomes when compared to a control group.

## Data Availability

All data produced in the present study are available upon reasonable request to the authors

