## Supplemental Table 1 for "Virtually Delivered Psychosocial Intervention for Mothers Expecting a Baby with Congenital Heart Disease: A Proof-of-Concept Study of HEARTPrep"

Supplementary Table 1. Content of HEARTPrep Modules

| **Module** | **Video Title** | **Video Description** | **Article Title** | **Article Description** |
| --- | --- | --- | --- | --- |
| Module 1: Adjusting | After the Diagnosis | Hear how other parents felt after the diagnosis. See how their children are doing now. | Your Feelings After the Diagnosis | It's normal and expected to have many different feelings after finding out your baby has a heart condition. |
|  | Preparing and Coping with Uncertainty | Hear how other parents prepared for their baby's birth. See how their children are doing now. | Ways to Handle Difficult Feelings | You might feel anxious, sad or angry at times. You might also feel hopeful. This is OK, and it's normal. |
| Module 2: Connecting | Navigating the Diagnosis as a Couple | Hear how other couples navigated the diagnosis. Learn about their relationships now. | Your Relationships After the Diagnosis | Your partner, your family and your friends can support you. But at times, these relationships can be challenging. |
|  | Navigating Relationships and Finding Support | Hear how other parents navigated relationships and found support after their baby's diagnosis. | Ways to Handle Difficulties in Relationships | When a relationship feels tense or you feel alone, there are things you can do. |
| Module 3: Preparing | Preparing for the Birth | Hear how other parents felt leading up to the birth and how they prepared. | Your Feelings Leading up to the Birth | It's normal to have a range of thoughts and feelings in the weeks leading up to your baby's birth. |
|  | Words of Hope and Advice | Hear words of hope and advice from other parents. See how their children and families are doing now. | Ways to Handle Anxiety | Feelings of anxiety or fear are normal leading up to your baby's birth and while your baby is in the hospital. |
